# Masculinizing Testosterone Therapy Reduces the Incidence of PIK3CA-Mutant/ER⁺ Breast Cancer but Not BRCA1-Associated Triple-Negative Breast Cancer

**DOI:** 10.1101/2025.09.15.25335324

**Authors:** Lin Wang, Brian R Sardella, Abhishek Thavamani, Erica S Massicott, Vanessa C Bret-Mounet, Gabrielle M Baker, Yaileen D Guzman-Arocho, Adam M. Tobias, Richard A. Bartlett, Emily K Aronson, Steven R Vandal, Zhaoji Liu, Jonah Lee, Mitko Veta, Suzanne C Wetstein, Sai Tun Hein Aung, Michelle L Lui, Kenrick Cato, Christine H Rohde, Kevin L Gardner, Hanina Hibshoosh, Walter O Bockting, Lauren C Houghton, Brittany M Charlton, Shana A Berwick, Alicia C Smart, Megan E Tesch, Arielle J Medford, Cornelia W Peterson, Jason D Domogauer, Li Jia, John G Clohessy, Nadine M Tung, Gerburg M Wulf, Yujing J Heng

**Affiliations:** Department of Medicine, Beth Israel Deaconess Medical Center & Harvard Medical School, Boston, MA, USA; Department of Pathology, Beth Israel Deaconess Medical Center & Harvard Medical School, Boston, MA, USA; Department of Surgery, Beth Israel Deaconess Medical Center & Harvard Medical School, Boston, MA, USA; Preclinical Murine Pharmacogenetics Facility and Mouse Hospital, Beth Israel Deaconess Medical Center, Boston, MA, USA; Medical Image Analysis Group, Eindhoven University of Technology, Eindhoven, the Netherlands; Columbia University Mailman School of Public Health, New York, NY, USA; Department of Pathology, Immunology & Laboratory Medicine, Rutgers New Jersey Medical School, Newark, NJ, USA; Department of Family and Community Health, University of Pennsylvania School of Nursing, Philadelphia, PA, USA; Division of Plastic and Reconstructive Surgery, Columbia University Irving Medical Center, New York, NY, USA; Department of Pathology and Cell Biology, Columbia University Irving Medical Center, New York, NY, USA; Department of Psychiatry and School of Nursing, Columbia University Irving Medical Center, New York, NY, USA; New York State Psychiatric Institute, New York, NY, USA; Columbia University Herbert Irving Comprehensive Cancer Center, New York, NY, USA; Department of Population Medicine, Harvard Pilgrim Health Care Institute & Harvard Medical School, Boston, MA; Department of Epidemiology, Harvard T.H. Chan School of Public Health, Boston, MA; The Fenway Institute, Boston, MA; Department of Radiation Oncology, Brigham and Women’s Hospital/Dana-Farber Cancer Institute & Harvard Medical School, Boston, MA, USA; Department of Radiation Oncology, Massachusetts General Hospital Cancer Center, Harvard Medical School, Boston, MA, USA; Department of Medical Oncology, Dana-Farber Cancer Institute & Harvard Medical School, Boston, MA, USA; Department of Medical, Massachusetts General Hospital Cancer Center & Harvard Medical School, Boston, MA, USA; Department of Comparative Pathobiology, Tufts University Cummings School of Veterinary Medicine, North Grafton, MA, USA; Department of Radiation Oncology, New York University Langone Health, New York University, New York, New York; Division of Urology, Department of Surgery, Brigham and Women’s Hospital & Harvard Medical School, Boston, MA, USA

**Keywords:** transgender health, gender-affirming hormones, transmen, breast cancer, breast cancer risk

## Abstract

**Background:** We investigated the impact of gender-affirming testosterone therapy (TT) on breast cancer (BC) risk and tumor progression.

**Materials and methods:** We leveraged a large human breast tissue dataset (n=417) to assess TT and terminal duct lobular unit (TDLU) involution, complemented with tissue markers (ER, PR, AR, and Ki67; *n*=24) and transcriptome profiling (*n*=8). Preclinical models assessed the effect of TT on BC incidence (*MMTV-Cre Pik3ca^f/wt^ n*=149 and *K14-Cre Brca^f/f^Tp53^f/f^ n*=153), murine mammary gland architecture (*n*=60), and tumor transcriptome (*n*=10). Lastly, we discuss trans masculine invasive BC cases and summarize tumor characteristics in this population (*n*=24).

**Results:** TT promotes TDLU involution by reducing epithelial proliferation via altered estrogen signaling and increases ER+, PR+, and Ki67+ extralobular stromal cells. In mice, TT similarly reduced mammary gland ductal branching and terminal end buds. TT decreased *Pik3ca*-related ER+ BC incidence by 81% compared to female controls (adj RR 0.19, 95% CI 0.08-0.45), but did not affect *Brca1*-related triple negative BC incidence. TT did not influence tumor progression in either model but shaped the *Pik3ca*-related ER+ tumor microenvironment toward a pro-tumor phenotype. Most trans masculine BC cases were ER+ (83.3%), small and node-negative, but were also moderately to poorly differentiated (70.8%).

**Conclusion:** TT reduces ER+ BC risk but does not eliminate risk, and has a negligible impact on *BRCA1*-related triple-negative BC risk. TT does not affect tumor growth once tumors are established but modulates the tumor microenvironment. Our work supports the need for breast cancer screening in TT users.

*Highlights:*

- TT reduces but does not completely ablate the breast epithelium.
- TT decreases *PIK3CA*-related ER+ breast cancer incidence by 81% compared to female control mice (adj RR 0.19, 95% CI 0.08-0.45), but does not affect *BRCA1*-related triple negative breast cancer incidence.
- TT does not affect tumor progression once the tumor is established.
- Trans masculine breast tumors are mostly ER+ (83.3%), small and node-negative, but are also moderately to poorly differentiated (70.8%).
- Tailored risk assessment and ongoing surveillance strategies are key for the care of transmasculine individuals who use TT.

## Introduction

Trans masculine individuals—transgender men and non-binary individuals assigned female at birth—may pursue gender-affirming testosterone therapy (TT) to treat their gender dysphoria ^1^. TT increases baseline testosterone levels by ≥10-fold to male physiological levels ^2^, which is 10 times higher than doses typically prescribed for hypoactive sexual desire disorder in cisgender women ^3^. The effects of TT on hormone-dependent malignancies such as breast cancer (BC) are not well understood.

The lifetime BC risk for females ranges between 8% and 12%, while it is 0.1% for males. BC risk increases to as high as 80% and 7% in female and male *BRCA1/2^mut^* carriers, respectively ^4,5^. The extent to which TT affects the BC risk of the general trans-masculine population and in high-risk *BRCA1/2^mut^* carriers is unclear. One epidemiologic study estimated the lifetime BC risk for trans masculine people to be 4% ^6^, which is lower than that of females but remains higher than that of males. In stark contrast, studies in pre- and postmenopausal women consistently link higher testosterone levels with increased BC risk ^7–9^. One explanation is that testosterone can be aromatized to estradiol, contributing to breast epithelium proliferation and tumorigenesis ^10^. Yet, evidence also exists that TT disrupts the homeostasis between estrogen and testosterone in the breast—our group showed that TT alters the histology and tissue composition of non-cancer breast tissue ^11–13^.

Chest-contouring surgeries differ from oncologic mastectomies because they focus on aesthetics and may leave significant residual breast tissue and muscle fascia. There is insufficient evidence to determine whether these procedures reduce risk. BC can occur after chest-contouring surgery ^14^. Furthermore, not all trans masculine individuals undergo chest-contouring surgeries, and while some may opt for puberty blockers, the majority will not and thus proceed through puberty ^15^. Lastly, the misunderstanding of their risk can lead to dismissal of symptoms or unwelcoming clinical spaces, therefore potentially delaying diagnosis ^16^. Therefore, understanding how TT influences BC incidence is crucial.

Terminal duct lobular unit (TDLU) involution in females is a natural phenomenon that occurs with aging ^17^, which results in a decrease in the number and/or size of the TDLUs, as well as a reduction in the number of acini within the lobules. Higher degrees of TDLU involution are linked to lower BC risk ^17^, hence TDLU involution is a potential surrogate marker for BC risk ^17–20^. Prospective human studies to understand the impact of TT on BC incidence will take decades. Even if available, the interpretation of human data will be complicated by noise due to many potential confounding factors. We sought to overcome those obstacles by taking a translational approach. We analyzed human breast tissues to understand TT and TDLU involution, complemented by tissue marker analysis and transcriptome profiling. As the hormone regulation of breast development is similar in mice and humans ^21^, we used *MMTV-Cre Pik3ca^f/wt^* and *K14-cre Brca1^f/f^Tp53^f/f^* mouse models to examine the effect of TT on *Pik3ca*-related estrogen receptor positive (ER+) and *Brca1*-related triple negative breast cancer (TNBC) incidence, respectively. As part of our correlative analyses, we present trans masculine invasive BC cases and summarize tumor characteristics in this population.

## Methods

### Human subjects and samples

Formalin-fixed paraffin-embedded (FFPE) breast tissue blocks were obtained from 444 subjects who had gender affirming chest-contouring surgery between 2013 and 2019 at Beth Israel Deaconess Medical Center (BIDMC) ^22^. These samples were used for TDLU involution and transcriptome profiling. This study was approved by the BIDMC Committee for Clinical Investigations (2018P000814).

Another set of 24 FFPE breast samples were obtained at Columbia University Irving Medical Center for immunohistochemistry (IHC) analysis—6 TT users who had gender affirming chest-contouring surgery between 2009 and 2020, 8 contralateral breasts of cisgender women with BC, and 10 adjacent normal chest tissue in cisgender men with gynecomastia. This study was approved by the Columbia University IRB (AAAT0129).

The Dana Farber/Harvard Cancer Center Office for Human Research Studies IRB determined that the medical records review of the four new invasive BC cases meets the criteria for exemption from IRB review (25-223).

### Animal experiments

All animal experiments were conducted in accordance with BIDMC IACUC protocol (#052-2020-23). Oophorectomy was performed at age 7 weeks. Mice slated for TT received weekly 200 ng of testosterone cypionate, starting at age 8 weeks. When a tumor is established, it was measured three times a week. The endpoint for each mouse was when the largest tumor reached 8 mm in longest diameter or, if multiple tumors arise, when the combined tumor burden reaches 20 mm. Mice with poor body condition or did not reach endpoint by age 58 weeks were euthanized and censored. We performed necropsy and counted the number of tumors in each mouse to assess tumor burden. Mammary tumors were confirmed by histology (GMW and CWP).

### TDLU involution assessed by pathologists and by automated method

Pathologists (GMB and YDG) were blinded to TT use and assessed TDLU involution as minimal (0–25%), mild (26–50%), moderate (51–75%), and marked (>75%) ^11^. TDLU involution was defined as an overall decrease in the number and size of the acini in each breast lobule. H&E slides (*n*=1073) from 417 subjects were digitized at 20× into whole slide images (WSIs) and processed by our previously developed image analysis method ^18,20^ (Fig. 1A). Our automated method captured four quantitative measures of TDLU involution: 1) number of TDLUs per mm^2^ of tissue area, 2) median TDLU size (mm^2^), 3) proportion of the breast tissue made up by TDLUs expressed as a percentage (i.e., % TDLU area), and 4) median number of acini per TDLU.

**Figure 1.**
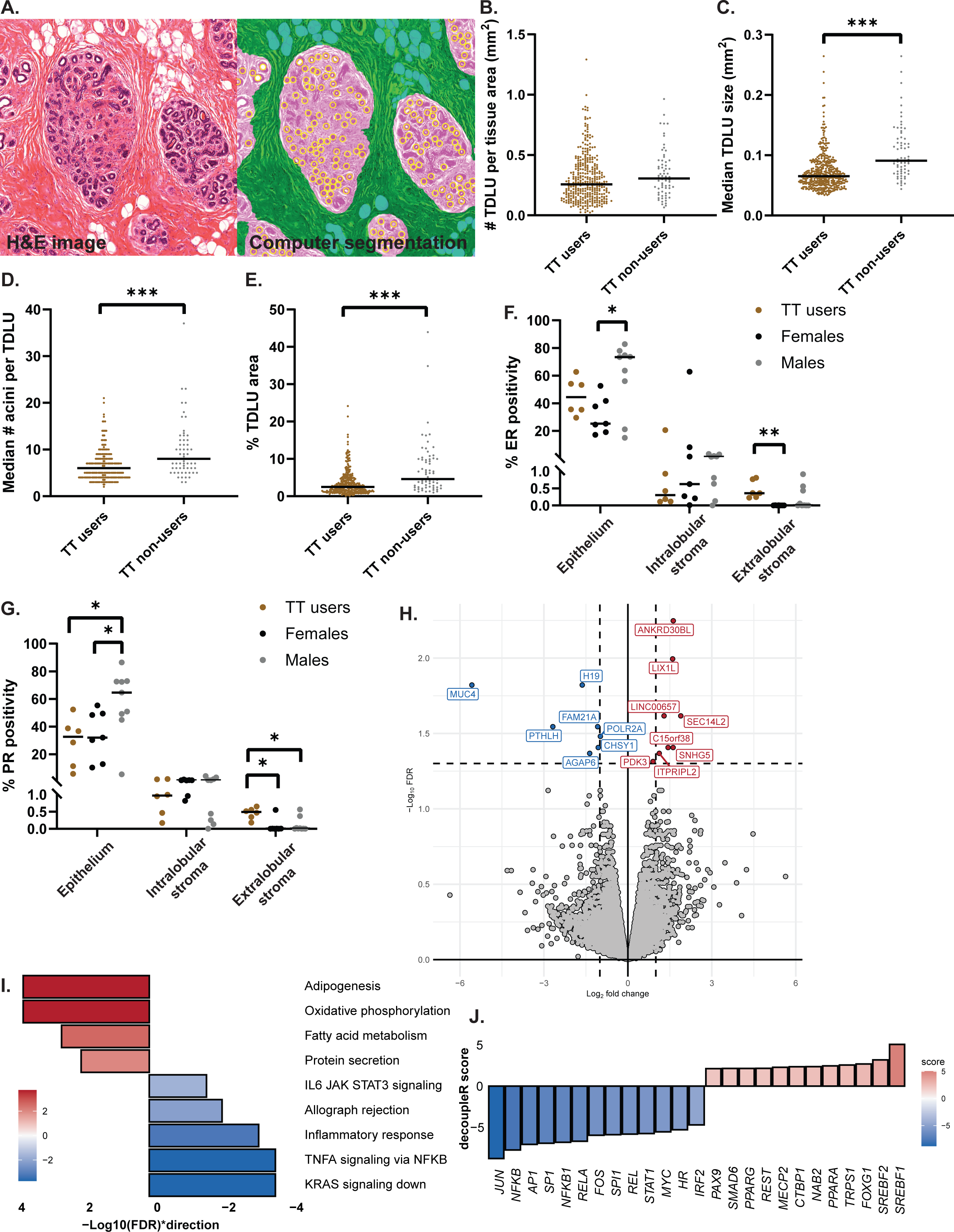
**A.** Our image analysis method segments an H&E breast whole slide image (left) into three components on the right panel: terminal duct lobular units (TDLU; pink areas), acini (yellow circles), and fat (blue areas; right). For illustration purpose, extralobular stroma in the right panel is colored green. The number of TDLUs per mm^2^ of tissue area captured by the automated method did not differ between testosterone therapy (TT) users and non-users (**B;** *p*=0.07). However, the size of TDLUs in TT users were smaller (**C;** *p*<0.001) and had less acini (**D;** *p*<0.001) compared to non-users, leading to lower %TDLU area (**E**; *p*<0.001). **F.** The percentage of estrogen receptor positive (%ER+) epithelial cells were significantly higher in cisgender males with gynecomastia than cisgender females (*p*=0.02) while TT users had more %ER+ stromal cells than females (*p*=0.004). **G.** The percentage of progesterone receptor positive (%PR+) epithelial cells were higher in cisgender males compared to TT users (*p*=0.04) and cisgender females (*p*=0.046). In contrast, %PR+ extralobular stromal cells were higher in TT users than cisgender females (*p*=0.01) and males (*p*=0.02). **H.** Volcano plot annotated with the 15 genes significantly differentially expressed between TT users and non-users (FDR<0.05). The red and blue dots represent genes that were up or down regulated, respectively. **I.** Significantly enriched Hallmark gene sets between TT users and non-users (FDR<0.05). Red and blue bars represent gene sets that were up or down regulated, respectively. **J.** Top 25 transcription factors activated (red) or deactivated (blue) in response to TT.

### Murine mammary gland architecture

Pairs of fourth mammary glands were harvested for whole mount analysis and were stained using the VitroView^™^ kit according to manufacturer’s instructions (VitroVivo Biotech, Rockville, MD). Slides were photographed, and branching and budding were analyzed using FIJI/ImageJ2 (version 2.14.0/1.54f).

### IHC and western blotting

IHC was performed using standard protocols and commercial antibodies. IHC quantification was performed using QuPath v0.6.0 by a pathology resident (SA) and a pathologist (HH). Western blotting adhered to the LI-CORbio^TM^ system protocol (Lincoln, NE).

### Transcriptome profiling

RNASeq was performed on FFPE breast tissue cores from four subjects who received TT between 15 and 17 months and four age-matched subjects who did not take TT, and spontaneous murine tumors (TT-treated *n*=5 and controls *n*=5). Data are available at the National Center for Biotechnology Information Gene Expression Omnibus (GSE306236).

### Statistical analysis

#### Human studies

Mann-Whitney or Fisher’s exact test were used to compare clinical characteristics. Correlations were performed using Spearman’s ρ. Linear regression evaluated the relationship between TT and each automated TDLU involution measure. TT non-users were assigned as zero months. Model 1 adjusted for age, year of surgery, and length of TT use (months). Model 2 additionally adjusted for race/ethnicity, BMI, whether the subject removed their ovaries at the time of chest-contouring surgery, and estimated weekly testosterone (mg). The percentages of IHC positive cells were compared using Kruskal-Wallis test and Dunn’s post hoc test.

#### Mouse studies

Log-rank test compared tumor-related survival between the study arms. Fisher’s exact test compared the proportion of tumor bearing mice. Adjusted relative risk (RR) was calculated using log-binomial mixed effect model (lme4 R package). Tumor doubling time was calculated using this formula: (*t* x log_10_(2))/(log_10_(V*_t_*/V*_0_*)), where V*_t_* equals the volume at time *t* and V*_0_* equals the volume at *t* = 0. One-way ANOVA with the appropriate post hoc test was used to compare between the groups. Paired *t*-test was used to compare the effect of *MMTV Cre*-recombinase on mammary gland branching and budding. Analyses were performed using R and GraphPad Prism v10.4.1. More experimental details and analysis are in Supplementary Methods.

## Results

### TT promotes TDLU involution

To understand the effect of TT on the architecture of the breast, we studied TDLU involution. Subjects with automated TDLU involution data were between 18 to 61 years old and 83.9% used TT (Table 1). Subjects tended to use TT for one to two years (39.7%) before undergoing chest-contouring surgery. TT users were also three years younger (*p*=0.003) and more likely to bind their chest (*p*=0.03) compared to non-users. Breast tissues of TT users were more likely to have moderate and marked TDLU involution compared to non-users when assessed by the pathologists (*p*<0.001; Table 1). Automated TDLU involution measures significantly correlated with pathologists’ assessment (all *p*<0.001), validating that our image analysis method was successfully applied to this new dataset (Suppl 1).

**Table 1.** Trans masculine subjects, stratified by testosterone therapy (TT).

|  | Used TT | Did not use TT | *p* value |
| --- | --- | --- | --- |
| N (%) | 350 (83.9) | 67 (16.1) |  |
| Age at surgery, median [IQR] | 24.0<br>[21.0, 29.0] | 27.0<br>[23.5, 32.0] | 0.003<sup>a</sup> |
| Race/ethnicity, *n* (%) |  |  | 0.04<sup>b</sup> |
| White | 274 (78.3) | 48 (71.6) |  |
| Black or African American | 28 (8.0) | 6 (9.0) |  |
| Asian | 19 (5.4) | 1 (1.5) |  |
| Multiracial | 18 (5.1) | 4 (6.0) |  |
| Native American/Pacific Islander | 1 (0.3) | 1 (1.5) |  |
| Unknown | 10 (2.9) | 7 (10.4) |  |
| BMI at surgery, median [IQR] | 25.7<br>[22.8, 29.7] | 25.1<br>[22.9, 29.4] | 0.58<sup>a</sup> |
| Chest binding, *n* (%) |  |  | 0.03<sup>b</sup> |
| Yes | 164 (46.9) | 20 (29.9) |  |
| No | 15 (4.3) | 6 (9.0) |  |
| Unknown | 171 (48.9) | 41 (61.2) |  |
| Duration of TT, *n* (%) |  |  | - |
| <1 year | 92 (26.3) | - |  |
| >=1 to <2 years | 139 (39.7) | - |  |
| >=2 to <5 years | 93 (26.6) | - |  |
| >=5 years | 12 (3.4) | - |  |
| Currently receives, unknown duration | 14 (4.0) | - |  |
| Oophorectomy prior to surgery, *n* (%) |  |  | 0.22<sup>b</sup> |
| Yes | 19 (5.4) | 1 (1.5) |  |
| No | 331 (94.6) | 66 (98.5) |  |
| TDLU involution (by pathologists), *n* (%) |  |  | <0.001<sup>b</sup> |
| Minimum | 61 (17.4) | 26 (38.8) |  |
| Mild | 96 (27.4) | 23 (34.3) |  |
| Moderate | 136 (38.9) | 16 (23.9) |  |
| Marked | 57 (16.3) | 2 (3.0) |  |

*p*-values were obtained using the <sup>a</sup>Mann-Whitney or <sup>b</sup>Fisher's exact test. Body mass index, BMI; TDLU, terminal duct lobular unit, inter-quartile range, IQR. Percentages may not add up to 100% due to rounding.

The number of TDLUs did not differ between TT users and non-users (Fig. 1B; *p*=0.07). However, TDLUs of TT users were 37% smaller (Fig. 1C; *p*<0.001), had 32% fewer acini (Fig. 1D; *p*<0.001), and correspondingly a 52% lower proportion of breast tissue was comprised of TDLUs compared to non-users. (Fig. 1E; *p*<0.001). These findings remained significant after adjusting for race/ethnicity, BMI, oophorectomy, and testosterone dose (Suppl 2).

### TT is associated with increased hormone receptor and Ki67 expression in extralobular stromal cells

As expected, ER and PR expression were highest in the epithelium, while typically <10% of the intralobular stroma and <1% of the extralobular stroma expressed ER or PR (Figs. 1F and 1G; Suppl 3 and 4). The percentage (%) of ER+ and PR+ epithelial cells was higher in cisgender males compared to cisgender females (1.9-fold higher for ER, *p*=0.02, and 1.7-fold higher for PR, *p*=0.046; Figs. 1F and 1G). While TT users had similar percentages of ER+ and PR+ epithelial cells as females, they exhibited a higher proportion of ER+ and PR+ cells in the extralobular stroma (p=0.004 and p=0.01), respectively; Figs. 1F and 1G). Androgen receptor (AR) expression did not differ between the groups (*p*>0.05; Suppl 5A). Proliferative activity throughout the breast was generally low with Ki67 <10% (Suppl 5B). TT users had more Ki67+ extralobular stromal cells than females (*p*<0.001) and males (*p*=0.04; Suppl 5B). Males also had more Ki67+ extralobular stromal cells than females (*p*=0.04; Suppl 5B). Our morphological and IHC data show that TT use leads to TDLU atrophy while increasing the hormonal responsiveness and proliferative capacity of the extralobular stroma.

### TT suppresses *MUC4* oncogene expression and KRAS signaling in the breast

To understand the effects of TT on the molecular mechanisms that drive TDLU involution, we performed bulk RNASeq on FFPE tissue cores of breast lobules. Consistent with our IHC data, *ER*, *PR*, and *Ki67* mRNA did not differ between TT users and non-users (Suppl 6 and 7A). We found 15 genes differentially expressed with TT use (Fig. 1H). The greatest difference was seen in *ANKRD30BL* (Ankyrin Repeat Domain 30B Like) expression, which was 3.1-fold higher in TT users (FDR=0.006; Suppl 7A). *ANKRD30BL* is most highly expressed in the testis ^23^. Thus, its upregulation in the breast tissue of in TT users is not unexpected although its role in the breast remains unclear ^24^. The expression of the cell surface glycoprotein and oncogene *MUC4* (Mucin 4) ^25^ was 98% reduced in TT users (FDR=0.02; Suppl 7A). Hallmark lipid metabolism-related gene sets were upregulated while KRAS signaling and immune-related gene sets were downregulated in TT users (all FDR<0.05; Fig. 1I). These findings are consistent with prior studies that show that androgens increase breast fatty acid metabolism ^26^ and can promote immunosuppression ^27^. Sex hormone gene sets were modulated in the expected directions although they did not reach significance—TT increased the androgen response (FDR=0.06) and downregulated the estrogen early and late responses (FDR>0.05; Suppl 7B).

We used decoupleR ^28^ to infer upstream regulatory activity driving transcriptional responses to TT. The top 25 transcription factors (Fig. 1J) generally agreed with our gene set enrichment results (Fig. 1I). The most activated transcription factor was *SREBF1* which maintains lipid homeostasis and supports breast milk production (Suppl 8) ^29^. In trans masculine people, the activation of *SREBF1* may explain the increased lipid metabolism caused by TT ^30^. Deactivated transcription factors were those involved in estrogen signaling (e.g. *JUN*, *AP1*, and *SP1*) and the immune response (e.g. *NFKB* and *SPI1*; Fig 1J). Consistently, single cell RNASeq in the human breast by Rath et al ^26^ showed that TT suppresses estrogen signaling by suppressing JUN and AP1 activity (Suppl 9). We did not observe increased amounts and activation of CD4+ T cells in TT-exposed breast tissues using EPIC ^31^, quanTIseq ^32^ (data not shown) or Gene Ontology Biological Processes signatures as reported by Rath et al ^26^. However, in our data, all 67 significantly enriched Gene Ontology Biological Processes signatures were downregulated in TT users, including immune responses (Suppl 7C), further confirming our Hallmark results (FDR<0.05, Suppl 7B). In summary, TT promotes expression patterns that antagonize carcinogenesis, such as upregulation of *ANKRD30BL* and downregulation of cancer-associated glycoprotein *MUC4*, and a shift away from proliferative signaling towards lipid metabolism, consistent with the known function of testosterone to oppose and regulate estrogen-mediated breast epithelial development.

### TT reduces *Pik3ca*-related ER+ BC risk

Based on our observations in human breast tissue, we hypothesized that TT would decrease the incidence of ER+ but not of ER-BC. We studied the impact of TT on BC incidence in *MMTV-Cre Pik3ca^f/wt^* mice that model human sporadic ER+ BC driven by a *PIK3CA^H1047R^* mutation ^33^. *MMTV-Cre Pik3ca^f/wt^* tumors express ER at early (Fig. 2A) and late stages (Fig. 2B), confirming that these tumors maintain ER expression throughout tumor progression. Similar to humans, ER expression in these murine tumors is highly variable.

**Figure 2.**
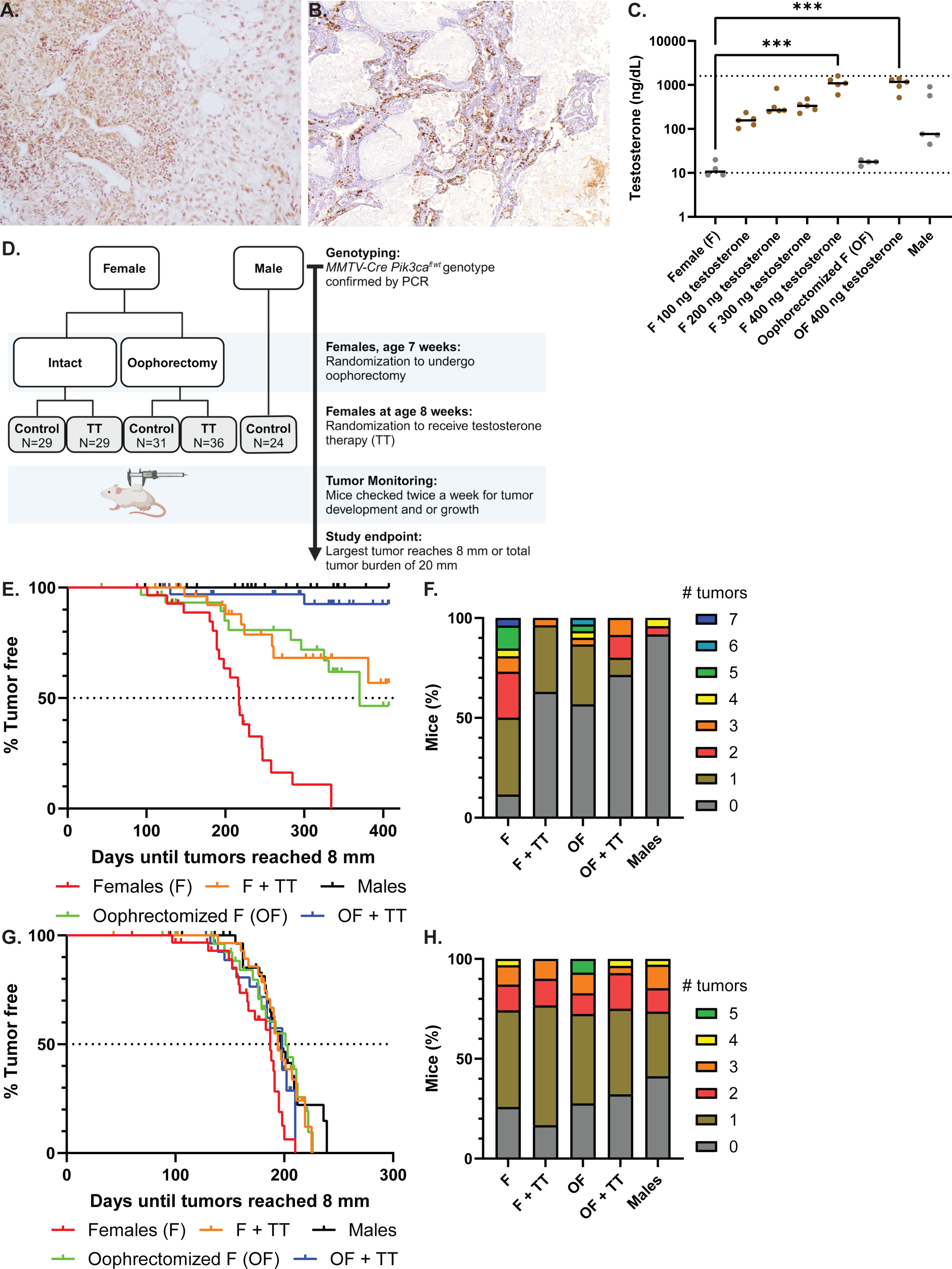
Examples of *MMTV-Cre Pik3ca^f/wt^* tumors expressing variable estrogen receptor (ER) immunohistochemistry expression at early (5-7 mm tumor burden; 20×; **A**) and at late stages (20 mm tumor burden; 10×; **B**). **C.** Serum testosterone levels at trough after four doses of testosterone showed that females and oophorectomized mice treated with 400 ng of weekly testosterone cypionate had significantly higher testosterone than untreated females (both *p*<0.001). The dotted horizontal lines represent the assay detection limits. **D.** Study design using *MMTV-Cre Pik3ca^f/wt^* mice to study the effect of testosterone therapy (TT) on *PIK3CA*-related, ER+ breast cancer incidence. **E.** Kaplan-Meier plot displaying the time taken for *MMTV-Cre Pik3ca^f/wt^* tumors to reach 8 mm. Female controls had the lowest median tumor-related survival of 217 days (red line). **F.** Stacked bar graphs showcase the proportion of mice (%) bearing between 0 and 7 *MMTV-Cre Pik3ca^f/wt^* tumors in each study arm. **G.** Time taken for *K14-Cre Brca1^f/f^Tp53^f/f^* tumors to reach 8 mm. Female controls had the lowest median survival of 187 days (red line). **H.** Proportion of mice (%) bearing between 0 and 5 *K14-Cre Brca1^f/f^Tp53^f/f^* tumors in each study arm.

Testosterone cypionate is used clinically ^22^ and in animal experiments ^34^. We determined that female mice treated with weekly 200 ng of testosterone cypionate achieved serum testosterone levels comparable to males ^2^ (Fig. 2C) and set up a five-arm experiment (*n*=149; Fig. 2D). Female controls developed breast tumors around age 5 months, with 50% reaching study endpoint of 8 mm tumor burden at age 217 days (Fig. 2E). Oophorectomized mice allow us to evaluate the effect of TT in the absence of endogenous estrogen and determine whether the aromatization of exogenous testosterone affects breast tumorigenesis ^35^. Oophorectomy arms are also clinically relevant as some trans masculine people undergo oophorectomy ^22^. When compared to female controls, oophorectomized mice had a significantly longer median tumor-related survival of 370 days (*p*<0.001; Fig. 2E). This confirmed that tumorigenesis in *MMTV-Cre Pik3ca^f/wt^* mice depends on endogenous estrogen. Males did not develop BC by the end of the experiment as expected (Fig. 2E).

TT-treated mice—both intact and oophorectomized—had a significantly longer median survival compared to female controls with median survival not reached during the study period of 407 days (both *p*<0.001; Fig. 2E). The median survival in TT-treated oophorectomized mice was longer than that of untreated oophorectomized mice (*p*=0.004; Fig. 2E) and TT-treated intact females (*p*=0.01; Fig. 2E). Tumor doubling time did not differ between the study arms (*p*>0.05, Suppl 10A).

Mice have 10 mammary glands. We considered each mammary gland as an independent sample and calculated the adjusted RR of developing a *Pik3ca*-related ER+ tumor. Female controls had the highest proportion of tumor-bearing mice (88.5%; *p*<0.001), with 49.9% of them developing at least two tumors (Fig. 2F). When compared to females, oophorectomized mice were 65% less likely to develop tumors (adj RR 0.35, 95% CI [0.16, 0.75]); TT-treated intact and oophorectomized mice had an even lower risk (81% and 77% lower, respectively); and male mice had the lowest risk (91% lower; Table 2A). Risk did not differ between TT-treated and untreated oophorectomized mice (adj RR 0.65, 95% CI [0.25, 1.70]) and between TT-treated intact and TT-treated oophorectomized mice (adj RR 1.22, 95% CI [0.48, 3.09]). TT-treated oophorectomized mice were more likely to develop tumors that did not reach the euthanasia endpoint of 8 mm by the end of the experiment at age 58 weeks. When compared to male mice, TT-treated mice had a 2.6-fold greater risk of developing tumors (adj RR 2.60, 95% CI [2.59, 2.61]) and TT-treated oophorectomized mice had a 3.1-fold greater risk (adj RR 3.05, 95% CI [3.05, 3.06]; Table 2A).

**Table 2.** Relative risk for tumorigenesis in each mouse model.

| **A. MMTV-Cre *Pik3ca*<sup>f/wt</sup>** | Proportion of tumors (n) | Adjusted RR* | 95% CI | p value |
| --- | --- | --- | --- | --- |
| Females (F) | 54/260 (20.8%) | Reference | - | - |
| F + TT | 12/270 (4.4%) | 0.19 | 0.08,0.45 | <0.001 |
| Oophorectomized F (OF) | 27/300 (9.0%) | 0.35 | 0.16,0.75 | 0.01 |
| OF + TT | 20/350 (5.7%) | 0.23 | 0.11,0.50 | <0.001 |
| Males | 6/240 (2.5%) | 0.09 | 0.03,0.27 | <0.001 |
| OF | 27/300 (9.0%) | Reference | - | - |
| OF + TT | 20/350 (5.7%) | 0.65 | 0.25,1.70 | 0.38 |
| F + TT | 12/270 (4.4%) | Reference | - | - |
| OF + TT | 20/350 (5.7%) | 1.22 | 0.48,3.09 | 0.68 |
| Males | 6/240 (2.5%) | Reference | - | - |
| F + TT | 12/270 (4.4%) | 2.60 | 2.59,2.61 | <0.001 |
| OF + TT | 20/350 (5.7%) | 3.05 | 3.05,3.06 | <0.001 |
| **B. K14-Cre *Brca1*<sup>f/f</sup> *Tp53*<sup>f/f</sup>** | Proportion of tumors (n) | Adjusted RR* | 95% CI | p value |
| F | 36/310 (11.6%) | Reference | - | - |
| F + TT | 35/300 (11.7%) | 1.00 | 0.62,1.62 | 0.99 |
| OF | 38/290 (13.1%) | 1.13 | 0.71,1.81 | 0.60 |
| OF + TT | 29/280 (10.4%) | 0.89 | 0.54,1.47 | 0.65 |
| Males | 35/340 (10.3%) | 0.89 | 0.55,1.43 | 0.62 |
| OF | 38/290 (13.1%) | Reference | - | - |
| OF + TT | 29/280 (10.4%) | 0.79 | 0.46,1.35 | 0.39 |
| F + TT | 35/300 (11.7%) | Reference | - | - |
| OF + TT | 29/280 (10.4%) | 0.89 | 0.56,1.41 | 0.62 |
| Males | 35/340 (10.3%) | Reference | - | - |
| F + TT | 35/300 (11.7%) | 1.13 | 0.72,1.77 | 0.59 |
| OF + TT | 29/280 (10.4%) | 1.01 | 0.63,1.61 | 0.98 |

\*Relative risk (RR) model controlled for mouse as random effect as each mouse contributed 10 mammary glands. Testosterone therapy, TT. Confidence interval, CI.

These data show that in this model system TT significantly lowered the risk of *PIK3CA*-related ER+ BC, independent of ovarian function. However, risk remained significantly higher than that of male mice, indicating that TT reduced but did not abolish BC risk.

### TT does not modulate *Brca1*-related TNBC incidence

Epidemiologic studies estimating trans masculine BC risk do not distinguish between ER+ and ER-disease due to limited sample sizes ^6,36^. To model a BC subtype that typically occurs at a young age, is genetically determined, and prevalent in men ^4^, we used *K14-Cre Brca1^f/f^Tp53^f/f^* mice ^37^ to investigate the effect of TT on *BRCA1-*related TNBC incidence (*n*=153; Suppl 11). When possible, tumors were histologically confirmed as breast carcinoma as these mice can develop other types of tumors ^38^. Female mice exclusively developed mammary adenocarcinomas as previously observed ^37^. Metaplastic mammary tumors—adenosquamous or squamous cell—were observed in all other study arms (Suppl 12).

Female controls had the shortest median survival of 187 days. The median survival was longer in TT-treated mice (194 days, *p*=0.005), oophorectomized mice (201 days, *p*=0.007), TT-treated oophorectomized mice (198 days, *p*=0.04), and male mice (197 days, *p*=0.004; Fig. 2G) when compared to female controls. There was no difference in median survival between TT-treated intact or TT-treated oophorectomized mice versus males (both *p*>0.05, Fig. 2G). However, oophorectomy and TT only extended the lag time to endpoint (8 mm) only by 7 to 14 days compared to female mice. While these findings achieved statistical significance, the effect size was small and the differences were less than two weeks. This is in stark contrast to the >150 days difference observed in *MMTV-Cre Pik3ca^f/wt^* mice (Fig. 2E). Tumor doubling time did not differ between the study arms (*p*>0.05, Suppl 10B).

Most tumor-bearing mice had a single tumor, and there was no difference in the proportion of tumor-bearing mice between the study groups. (*p*=0.30, Fig. 2H). Neither oophorectomy nor TT affected the risk of developing a *K14-Cre Brca1^f/f^Tp53^f/f^* tumor compared to female or male mice (all *p*>0.05, Table 2B). *K14-Cre Brca1^f/f^Tp53^f/f^* tumorigenesis does not appear to be affected by estrogen or TT, and consistently these tumors have lower Ar protein expression than *MMTV-Cre Pik3ca^f/wt^* tumors (Suppl 13).

### TT reduces murine mammary gland branching and terminal end buds

Since TT decreased *MMTV-Cre Pik3ca^f/wt^* tumorigenesis strongly but not in *K14-Cre Brca1^f/f^Tp53^f/f^*, and given our observations of the effect of TT in the human breast (Fig. 1), we investigated how TT affected the structure of murine mammary glands. In all study arms, the number of branches and terminal end buds was significantly reduced when compared to female controls (all *p*<0.001, Figs. 3A to 3C). However, TT-treated females still exhibited a significantly greater degree of branching and budding compared to males (4.2-fold *p*=0.04 and 4.5-fold *p*=0.03, respectively; Figs. 3B and 3C). Similarly, TT-treated oophorectomized mice also had significantly more branching (3.9-fold, *p*=0.04) compared to males (Fig 3B). As a pertinent control, we confirmed that MMTV-driven Cre-recombinase alone did not affect branching or budding (both *p*=0.12; Suppl 14).

**Figure 3.**
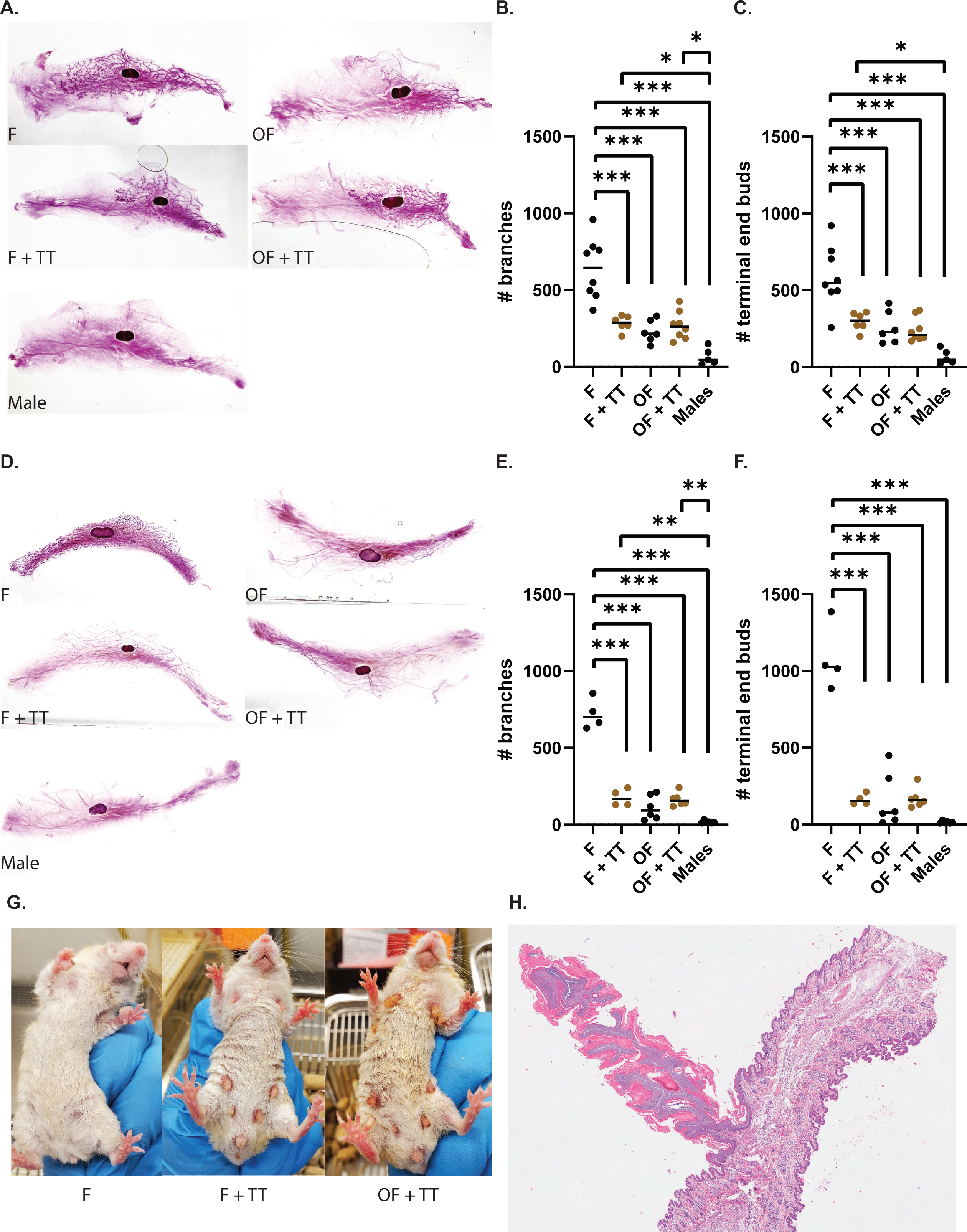
**A.** Examples of *MMTV-Cre Pik3ca^f/wt^* mammary gland whole mounts from each study arm. The number of mammary gland branching (**B**) and terminal end buds (**C**) were significantly lower in testosterone therapy (TT)-treated females, oophorectomized females (OF), TT-treated OF, and male mice compared to females (F). Similarly, *K14-Cre Brca1^f/f^Tp53^f/f^* mammary gland whole mounts (**D**) were assessed for branching (**E**) and terminal end buds (**F**). **G.** Examples of *MMTV-Cre Pik3ca^f/wt^* mice with nipple hypertrophy after 12 weeks of TT. **H.** A representative H&E section of a nipple from a TT-treated oophorectomized mice at 1.8×. There are scattered dyskeratotic keratinocytes in the nipple epidermis. Focal ductal structures and mammary epithelial elements in the underlying dermis are also present.

Similar findings were observed in *K14-Cre Brca1^f/f^Tp53^f/f^* mice where females had the most branching and budding compared to other study arms (all *p*<0.001, Figs. 3D to 3F). TT-treated females and oophorectomized mice had significantly more branching than males (10.6-fold *p*=0.005 and 9.8-fold *p*=0.004, respectively, Fig. 3E). There was no difference in branching and budding between TT-treated females, oophorectomized mice, and TT-treated oophorectomized mice in both models (all *p*>0.05).

### TT induces nipple hypertrophy in *MMTV-Cre Pik3ca^f/wt^* mice

While conducting our tumor-related survival studies, we observed nipple hypertrophy in *MMTV-Cre Pik3ca^f/wt^* mice treated with TT (Fig. 3G) but not in *K14-Cre Brca1^f/f^Tp53^f/f^* mice. Nipple hypertrophy typically manifests after four weeks of TT. Histopathologic review of these nipples revealed extensive hyperkeratosis with occasional focal parakeratosis and engorged blood vessels in the underlying dermis (Fig. 3H).

### TT modulates the *MMTV-Cre Pik3ca^f/wt^* tumor microenvironment

We performed RNASeq on tumors that arose from TT-treated females and female controls to elucidate the biology of TT-exposed tumors (Fig. 2E). Twelve genes were differentially expressed at *p*<0.001 (Suppl 7D). Six of those genes— *Nptx2, Gvin2, Pigr, Blnk, Gdpd3*, and *Mmd*—were immune-related genes upregulated in TT-exposed tumors (Fig. 4A). Thirty-three Hallmark gene sets were enriched in TT-exposed tumors (all FDR<0.05, Suppl 7E). TT-exposed tumors were more proliferative (*n*=9 gene sets) and exhibited a greater degree of inflammation (*n*=5 gene sets), and had a higher degree of glycolysis and oxidative phosphorylation (*n*=12 gene sets; Fig. 4B). Consistent with our human data in Fig. 1I, TT suppressed Kras signaling in these murine tumors (Fig. 4B). While *Ar* mRNA did not differ between TT-treated and control tumors (*p*=0.11; Suppl 7D), ANDROGEN RESPONSE and ESTROGEN RESPONSE (EARLY) gene sets were both upregulated in TT-exposed tumors (FDR<0.001 and FDR=0.01, respectively; Fig 4B).

**Figure 4.**
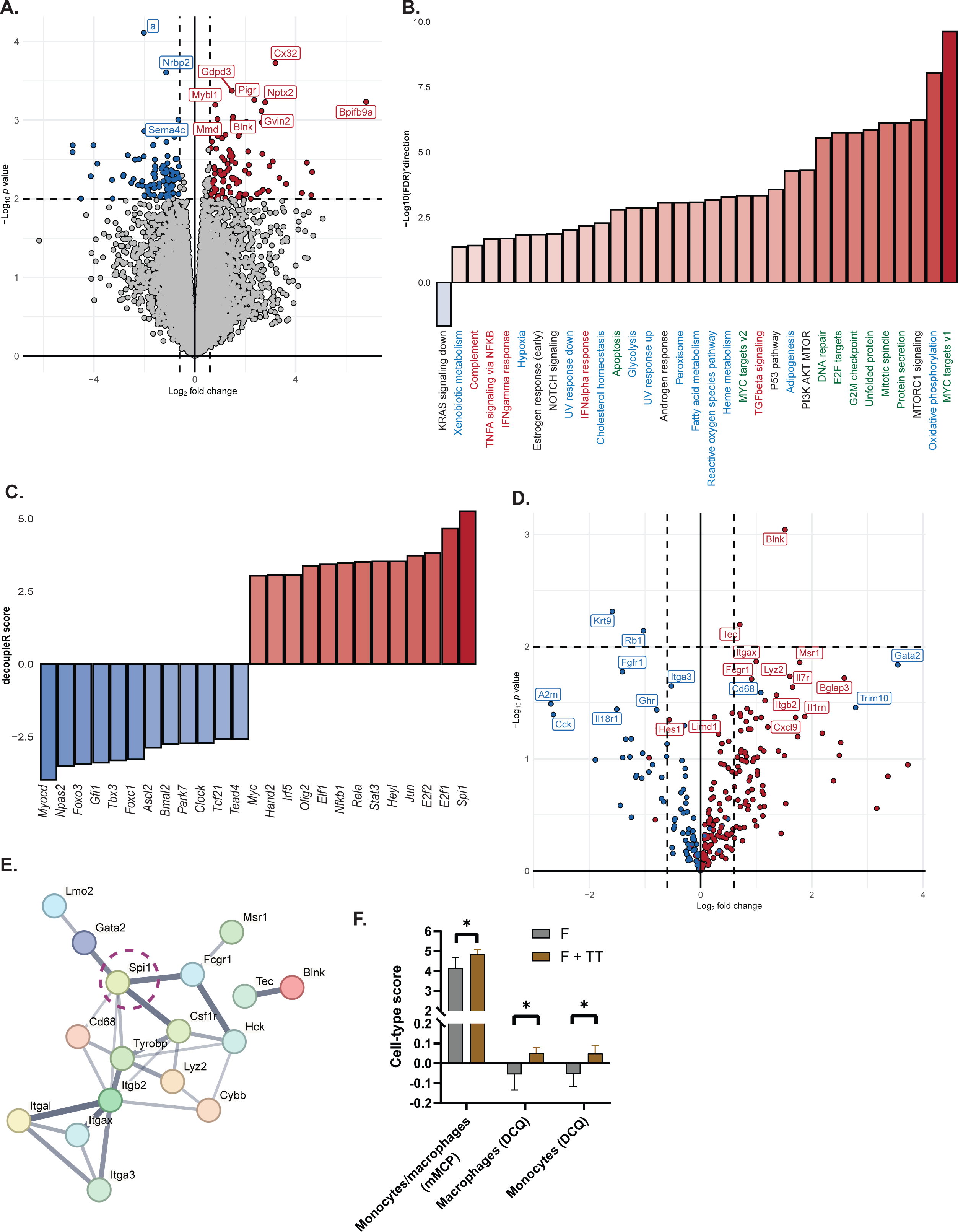
**A.** Twelve genes were differentially expressed between testosterone therapy (TT)-treated female *MMTV-Cre Pik3ca^f/wt^* tumors compared to female controls at *p*<0.001. The red and blue dots represent genes that were up or down regulated, respectively. **B.** Thirty-three Hallmark gene sets were significantly enriched in TT-treated tumors (FDR<0.05; red bars are upregulated gene sets; the blue bar shows a downregulated gene set). Gene sets related to inflammation are labeled with red text, proliferation with green text, and metabolism with blue text. **C.** Top 25 transcription factors predicted by decoupleR that were activated (red) and deactivated (blue) by TT. **D.** *Spi1* was predicted as the most activated transcription factor modulated by TT because the majority of its downstream target genes were upregulated (red). **E.** A protein-protein interaction network of Spi1. The line thickness represents the strength of data support. **F.** Two deconvolution methods—murine Microenvironment Cell Population Counter (mMCP) and digital cell quantification (DCQ) estimated an increase in monocytes and macrophages in TT-treated tumors compared to control (both *p*<0.05 and FDR<0.17).

The most activated transcription factor was *Spi1* (Figs. 4C and 4D). *Spi1* is a master regulator in myeloid and lymphocyte development ^39^. A protein–protein interaction network centered on Spi1 revealed that interacting proteins are associated with myeloid lineage differentiation, macrophage function, and immune signaling (Fig. 4E). The activation of *Spi1* is likely related to the enrichment of inflammatory genes and gene sets (Figs. 4A and 4B). *Ar* was neither activated nor deactivated in TT-exposed tumors (*p*=0.38) which was in line with our tumor-related survival findings (Fig. 2E and Suppl 10A) where TT did not affect tumor growth once the tumor was established.

The RNA-seq analyses indicate that TT modulates the tumor microenvironment. We estimated the tumor immune-cell composition using four murine-based deconvolution methods ^40^. The enrichment of immune gene sets in TT-exposed tumors may be the consequence of higher infiltration of TT-exposed tumors with monocytes and macrophages as shown by both murine Microenvironment Cell Population Counter (mMCP-counter) and digital cell quantification (DCQ) methods (all *p*<0.05 and FDR<0.17; Fig. 4F and Suppl 15). Androgen skews macrophage function toward an M2-like, pro-tumor phenotype ^41,42^. Androgen also drives CD8 T cell exhaustion in cancer ^27^. Gene signatures of CD8 progenitor exhaustion, effector-like, and terminal exhaustion ^43^ were all significantly upregulated in TT-exposed tumors compared to control using the ROAST method ^44^ (all *p*<0.001).

Research-based Oncotype Dx (21-genes) and MammaPrint (70-genes) scores did not differ between TT-exposed and control tumors (both *p*>0.05; Suppl 16). Except for one tumor, all tumors were classified as ‘high risk’ of recurrence with both signatures.

### Trans masculine invasive BCs

We present four trans masculine invasive BC cases in Suppl 17 and combined the clinical and tumor characteristics with 20 published cases to understand whether TT affects tumor aggressiveness (Suppl 18). Trans masculine patients were diagnosed at a relatively young age (median 42.8 years) and were taking TT between 3 months to 25 years at the time of diagnosis (median 3 years; Table 3). Over half of the tumors (13/23, 54.2%) were detected in intact breast tissue prior to top surgery, four cases (16.7%) were incidental findings in post-surgical specimens, and seven cases (29.2%) were identified in residual chest tissue between one and 12 years following top surgery (Table 3). A family history of cancer was reported in nine patients (37.5%; Table 3). Among the 11 individuals who underwent germline genetic testing, only one carried a pathogenic mutation which is related to melanoma and renal cell cancer but not BC (*MITF* c.952G>A; Suppl 16).

**Table 3.**
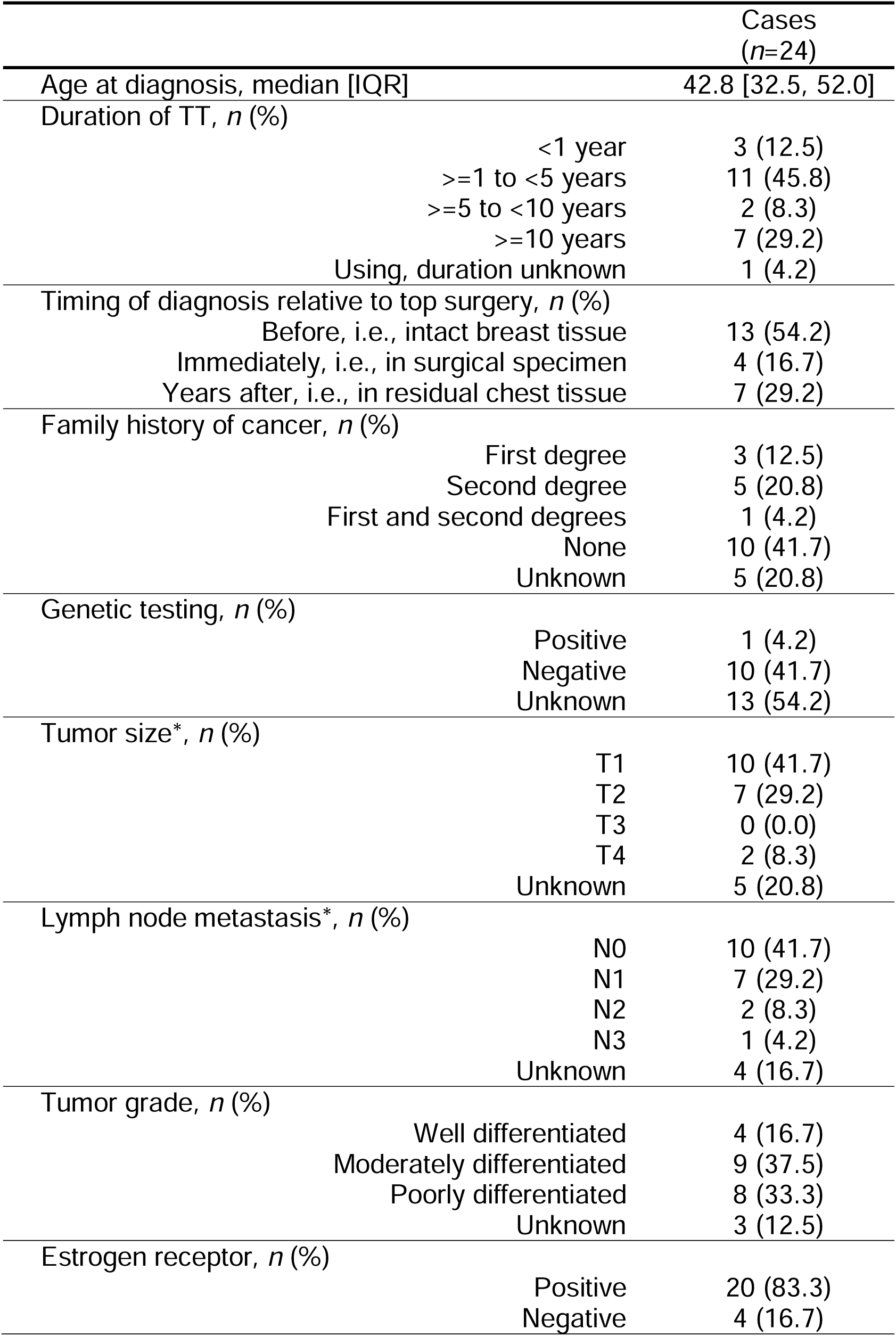
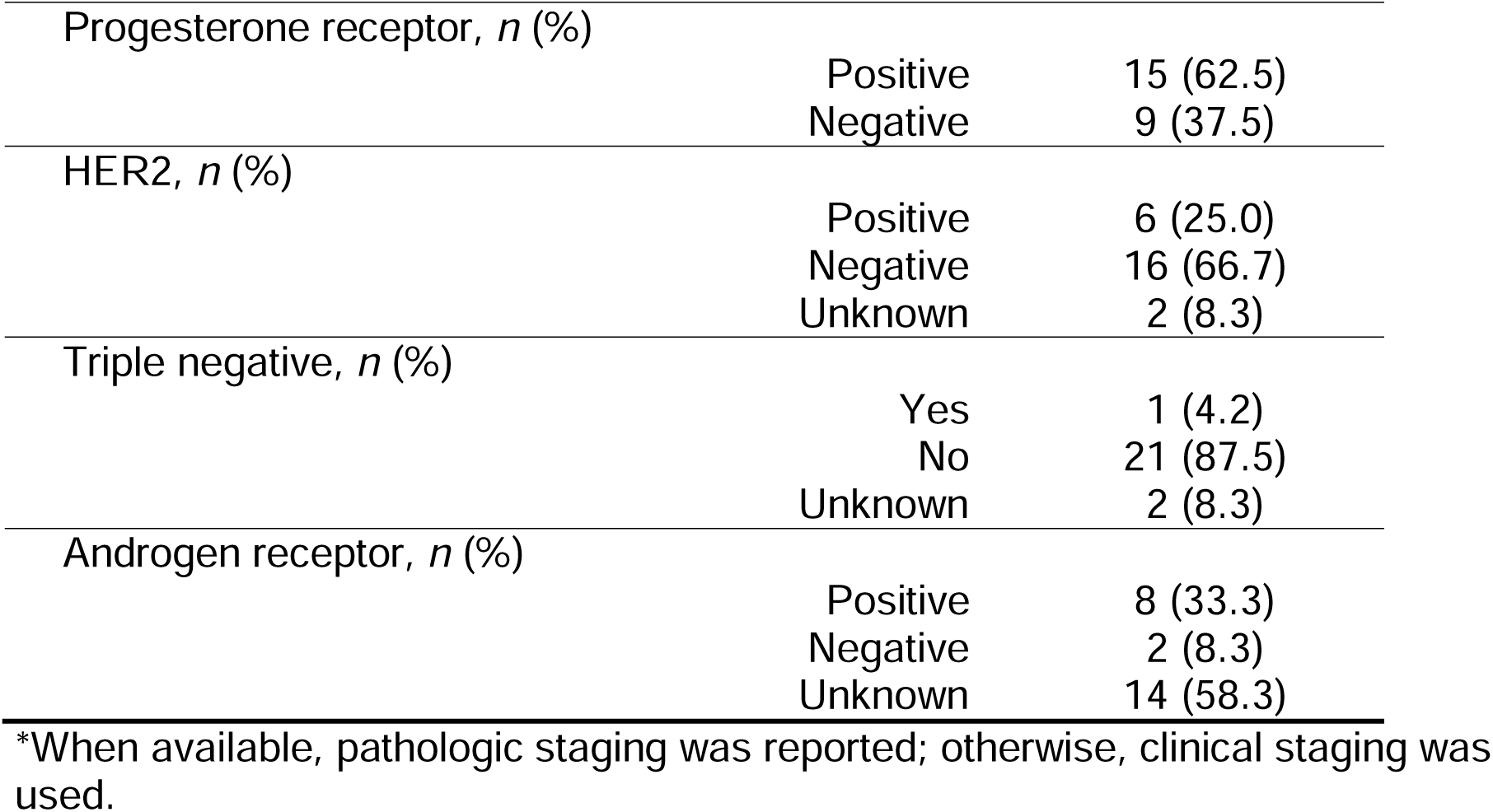
Clinical and tumor characteristics of invasive breast cancer cases in trans masculine people taking testosterone therapy (TT) at time of diagnosis.

|  | Cases<br>(*n*=24) |
| --- | --- |
| Age at diagnosis, median [IQR] | 42.8 [32.5, 52.0] |
| Duration of TT, *n* (%) |  |
| <1 year | 3 (12.5) |
| >=1 to <5 years | 11 (45.8) |
| >=5 to <10 years | 2 (8.3) |
| >=10 years | 7 (29.2) |
| Using, duration unknown | 1 (4.2) |
| Timing of diagnosis relative to top surgery, *n* (%) |  |
| Before, i.e., intact breast tissue | 13 (54.2) |
| Immediately, i.e., in surgical specimen | 4 (16.7) |
| Years after, i.e., in residual chest tissue | 7 (29.2) |
| Family history of cancer, *n* (%) |  |
| First degree | 3 (12.5) |
| Second degree | 5 (20.8) |
| First and second degrees | 1 (4.2) |
| None | 10 (41.7) |
| Unknown | 5 (20.8) |
| Genetic testing, *n* (%) |  |
| Positive | 1 (4.2) |
| Negative | 10 (41.7) |
| Unknown | 13 (54.2) |
| Tumor size*, *n* (%) |  |
| T1 | 10 (41.7) |
| T2 | 7 (29.2) |
| T3 | 0 (0.0) |
| T4 | 2 (8.3) |
| Unknown | 5 (20.8) |
| Lymph node metastasis*, *n* (%) |  |
| N0 | 10 (41.7) |
| N1 | 7 (29.2) |
| N2 | 2 (8.3) |
| N3 | 1 (4.2) |
| Unknown | 4 (16.7) |
| Tumor grade, *n* (%) |  |
| Well differentiated | 4 (16.7) |
| Moderately differentiated | 9 (37.5) |
| Poorly differentiated | 8 (33.3) |
| Unknown | 3 (12.5) |
| Estrogen receptor, *n* (%) |  |
| Positive | 20 (83.3) |
| Negative | 4 (16.7) |

|  |  |  |
| --- | --- | --- |
| Progesterone receptor, *n* (%) | Positive | 15 (62.5) |
|  | Negative | 9 (37.5) |
| HER2, *n* (%) | Positive | 6 (25.0) |
|  | Negative | 16 (66.7) |
|  | Unknown | 2 (8.3) |
| Triple negative, *n* (%) | Yes | 1 (4.2) |
|  | No | 21 (87.5) |
|  | Unknown | 2 (8.3) |
| Androgen receptor, *n* (%) | Positive | 8 (33.3) |
|  | Negative | 2 (8.3) |
|  | Unknown | 14 (58.3) |

\*When available, pathologic staging was reported; otherwise, clinical staging was used.

TT-exposed tumors were mostly ER+ (20/24, 83.3%), small and node-negative, but were moderately to poorly differentiated (17/24, 70.8%; Table 3). AR staining was performed in 10 cases, of which eight cases were AR+ (33.3%). Four cases had Oncotype Dx scores, ranging from 14 to 22, indicating a low risk of recurrence (Suppl 16).

## Discussion

TT use is expected to increase as more individuals identify as gender diverse. In the US, over 1.6 million people identify as transgender ^44^, with youth representing the fastest growing subgroup within the LGBTQ+ community ^44^. Similar trends are observed globally ^45–48^. As the young transgender population ages, it is important to understand the long-term health effects of gender-affirming hormone therapy and collect evidence to guide clinicians on cancer risks and surveillance in this population. We integrated human breast tissues and cancer case analyses with preclinical models to comprehensively analyze the impact of TT on BC risk.

TT promotes age-independent involution of TDLUs, reduces epithelial proliferation via altered estrogen signaling, and decreases oncogenic pathways in the breast. Our TDLU involution results are consistent with our prior work where TT users had 28% less epithelium compared to non-users when assessed using a different image analysis method ^13^. ER, PR and AR remain strongly expressed in the tissues of TT users ^45^. Essentially, the breast epithelium becomes rarefied while its composition remains similar to that of males and females. This reduction in ‘at-risk’ epithelium might explain why TT decreases but does not completely eliminate BC risk. The higher percentage of ER+, PR+, and Ki67+ extralobular stromal cells in TT users suggests an unrecognized role of fibroblasts in regulating TDLU involution. Our preclinical data suggest that TT may reduce the risk of ER+ BC by more than 50%. In our *Pik3ca*-related ER+ study, we demonstrated that the effects of lowered estradiol levels via oophorectomy and exogenous testosterone on ER+ tumorigenesis were not redundant but additive and may differ mechanistically. Our *Pik3ca*-related ER+ study coupled with our whole mount analysis support epidemiologic findings ^6,36^ that BC risk, mostly referring to ER+ risk, in trans masculine people is lower than that of cisgender women but remains higher than that of cisgender men.

The observation of nipple hypertrophy in *MMTV-Cre Pik3ca^f/wt^* mice was unexpected as *MMTV-Cre* is generally only active in the mammary epithelium. It appears that the hyperkeratosis is secondary to the activation of *Pik3ca* in the ductal cells in the nipple, in combination with direct testosterone effects on the skin. This is of interest because testosterone affects skin health in humans. TT users experience thicker and oilier skin ^2^ while cisgender men on testosterone replacement therapy experience nipple changes and tenderness ^46^. We reported that AR expressing Toker cell hyperplasia in the nipple-areolar complex is more common in trans masculine TT users compared to cisgender women ^12^.

Surprisingly, TT did not affect the incidence of *Brca1*-related TNBC. However, TT shifted the histological subtype from adenocarcinoma to a higher percentage of metaplastic tumors in TT-treated arms and male mice. Metaplastic BC are rare, hard to treat, and more frequently seen in patients with a hereditary BC risk ^47^. Although it remains uncertain whether this observation in mice will translate to humans, it raises the possibility that TT users may develop rarer BC subtypes at a higher frequency— a question that could be addressed using cancer registries and real-world databases as human data accumulate.

Over 80% of trans masculine invasive BC cases were ER+, which is slightly higher than in cisgender women (∼75%) ^48^, despite the trans masculine patients being diagnosed at a younger age. This mirrors cases in cisgender men where ∼90% of tumors are ER+ ^49^. The absence of TNBC may be explained by the atrophy of the basal layer of the TDLU ductal epithelium induced by TT. This, however, does not apply to the *BRCA1^mut^* setting as those tumors arise from uncommitted ER-/AR-luminal progenitor cells in the TDLU ^50^. Accordingly, TT did not affect tumor incidence in our *Brca1-*related TNBC mouse model. Clinical data regarding the effect of hormonal manipulations on BC risk in *BRCA1/2^mut^* carriers such as oophorectomy have shown disparate results ^51–53^. Our preclinical data suggest that *BRCA1/2^mut^* carriers who pursue TT are likely to retain their inherent risk of developing TNBC.

Given an approximate lifetime BC risk of 12% in cisgender women, our preclinical findings support the previously estimated 4% lifetime BC risk in TT users ^6^. Notably, this 4% risk is comparable to the lifetime colon cancer risk in the general population ^54^ and therefore warrants screening. The occurrence of BC both before and long after top surgery underscores the persistent risk of malignancy in residual chest tissue and highlights the need for nuanced risk assessment and ongoing screening strategies tailored to this population, irrespective of organ inventory, genetic predisposition, or duration of TT use. Our findings may reassure clinicians that prescribing TT is not associated with increased BC risk and could help reduce stigma surrounding its use in trans masculine individuals. Additionally, neither estradiol nor testosterone affected tumor progression once a tumor was established suggesting that gender-confirming hormones may not affect tumor growth after a diagnosis of BC.

Molecular findings in *Pik3ca*-related ER+ murine tumors further confirmed that TT had minimal impact on tumor progression and recurrence scores. Nevertheless, TT induced architectural and molecular changes in the tumor. In agreement with others ^27,41,42,55^, we showed that TT shapes the tumor microenvironment by modulating immune responses in macrophages and T cells towards a pro-tumor phenotype, while tumor-intrinsically, TT downregulated KRAS signaling in both human breast tissues and ER+ murine tumors, consistent with AR agonists acting as tumor suppressors in ER+ BC ^56,57^.

In conclusion, TT rarefies, but does not ablate the breast epithelium and reduces BC risk through TDLU atrophy and down regulation of oncogenic signaling. TT may promote immune evasion but does not affect tumor cell intrinsic features or tumor growth in *PIK3CA*-related ER+ disease.

## Notes: The role of the funder

The funding sources listed in the Funding section were not involved in the design of the study; the collection, analysis, and interpretation of the data; the writing of the manuscript; and the decision to submit the manuscript for publication.

## Acknowledgements

The authors assume full responsibility for analyses and interpretation of these data. Animal histology was performed at the BIDMC histology core facility (RRID:SCR_009669). Human breast RNASeq was performed at the Molecular Biology Core Facilities at Dana Farber Cancer institute, Boston, MA.

## Data Availability

The data that support the findings of this study are available from Dr. Jan Heng.

## Declaration of Generative AI and AI-assisted technologies in the writing process

During the preparation of this work the author(s) used ChatGPT to improve readability and language. After using this tool/service, the author(s) reviewed and edited the content as needed and take(s) full responsibility for the content of the publication.

